# The association between smoking and anal human papillomavirus in the HPV infection in Men Study

**DOI:** 10.1101/2021.10.18.21265161

**Authors:** Victoria Umutoni, Matthew B. Schabath, Alan G. Nyitray, Timothy Wilkin, Luisa Villa, Eduardo Lazcano-Ponce, Anna R. Giuliano, Staci L. Sudenga

## Abstract

**Background:** Previous studies show an association between smoking and anal cancer. The objective of this study was to assess the association between smoking and anal HPV prevalence in men.

**Methods:** The HPV infection in Men (HIM) Study is a multinational study that enrolled HIV-negative men. At baseline anal specimens were collected from 1994 participants. HPV genotyping was assessed by Linear Array. Prevalence ratios (PR) were used to assess the association between smoking and anal HPV prevalence.

**Results:** Current smokers have a higher prevalence of any anal HPV (adjusted PR (aPR)=1.36, 95%CI: 1.06-1.73) and LR-HPV (aPR=1.59, 95%CI: 1.20-2.12) compared to never smokers. There were no difference in the prevalence of anal HPV between former and never smokers.

Smoking status was not associated with the prevalence of anal HPV among men that have sex with men (MSM). Among men that have sex with women (MSW), current smokers had an increased prevalence for LR-HPV (aPR=1.60 95% CI: 1.02-2.50) compared to never smokers.

**Conclusions:** While there was no difference in anal HPV prevalence among MSM by smoking status, MSW that currently smoked had a higher prevalence of LR-HPV. Futher research is needed to assess the role smoking in anal HPV persistence.

## Introduction

Anal cancer incidence has been shown to be increasing over time in both men and women from high resource countries, especially in those with immunocompromised conditions such as HIV.^1^ In the US, anal cancer incidence has increased 2.7% per year between 2001 to 2015 with increases in both men (2.2%) and women (3.1%).^2^ According to the Center for Disease Control, each year in the US there are 4700 new cases of HPV-associated anal cancers in women and 2300 in men.^3^

Risk factors for anal cancer includes human papillomavirus (HPV) infection, tobacco smoking, HIV infection, and a high number of sexual partners.^4^ Worldwide, 88% of anal carcinoma are caused by high-risk HPV ^5^ and 79% of those are attributable to HPV16 and HPV 18.^1, 6^

Previous studies have shown an association between smoking and HPV infection. ^7^ Smoking at least one pack year of cigarettes was associated with detection of anal HPV DNA.^8^ Smoking suppresses local immune function, increases cellular proliferation and turnover, upregulates proinflammatory factors, and can induce host DNA damage which could increase the likelihood of HPV persisting. ^7^ Studies among women have shown that cigarette smoking is associated with HPV infection and a higher HPV 16 and HPV18 viral load in current smokers compared to never smokers.^9^ We have previously shown that smoking was associated with an increased prevalence of genital HPV among men.^10^ The objective of this study was to assess the association between tobacco smoking and the prevalence of anal HPV among men residing in Tampa, Florida, US; Cuernavaca, Mexico; and Sao Paulo, Brazil. Analyses were stratified by sexual orientation given that anal HPV prevalence varies by sexual orientation.

## Methods

### Study Population

The HPV infection in men (HIM) study enrolled 4123 HIV-negative men aged 18-70 years and residing in Tampa, Florida, US; Cuernavaca, Mexico; and Sao Paulo, Brazil between July 2005 and June 2009. A full description of the study procedures has been published.^11^ At baseline participants were given a physical exam where multiple specimens were obtained for laboratory analysis and a self administered questionnaire looking at sociodemographic characteristics, sexual history, condom use, alcohol and tobacco use.

All participants provided written informed consent. Study protocols were approved by the Institutional Review Boards at the University of South Florida (Tampa, FL, US), the Ludwig Institute for Cancer Research, the Centro de Referencia e Treinamento em Doencas Sexualmente Transmissiveis e AIDS (São Paulo, Brazil), and the Instituto Nacional de Salud Pública (Cuernavaca, Mexico).

### Anal Specimen collection for HPV detection

Participants underwent a clinical examination at each visit. Using a pre-wetted Dacron swab, 360 degrees of the anal canal epithelium was swabbed between the anal opening and the anal canal dentate line, after which the swab was placed into standard transport medium and stored at at -80°C.^12^ Specimens underwent DNA extraction (Qiagen Media Kit), PCR analysis, and HPV genotyping (Roche Linear Array).^13^ If samples tested positive for beta-globin or an HPV genotype, they were considered adequate and were included in the analysis. The Linear Array assay tests for 37 HPV types, classified as high-risk (HR-HPV; types 16, 18, 31, 33, 35, 39, 45,51, 52, 56, 58, 59, and 68) or low-risk (LR-HPV; types 6, 11, 26, 40, 42, 53, 54, 55, 61, 62, 64, 66, 67, 69, 70, 71, 72, 73,81, 82, 82 subtype IS39, 83, 84, and 89).^14^ The HPV genotypes were further classified as the HPV types in the 4vHPV vaccine (6/11/16/18) and the 9vHPV vaccine (6/11/16/18/31/33/45/52/58).

### Statistical analysis

For this analysis, partipants who had an anal specimen (n=2030) collected were included. Participants with missing smoking data (n=39) were excluded. Participants self-reported the number of lifetime sexual partners at baseline and sex was defined as oral, vaginal or anal sex. Based on responses to these questions, sexual orientation was categorized into men who have sex with women (MSW) only and men who have sex with men irrespective of sex with women (MSM). Men that did not report any sexual partners at baseline were classified as unknown for sexual orientation. Tobacco smoking status was categorized as: never smoker, former smoker and current smoker. Smoking pack-years was calculated by taking number of packs smoked per year (number of cigarettes per day divided by 20 cigarettes per pack) multiplied by the number of years smoked. The median pack-years smoked was assessed. For current and former smokers seperately, tertiles for “number of cigarrettes smoked per day,” “number of years smoked,” and “smoking pack-years” were calculated.

The outcome of interest was prevalence of: any HPV, LR-HPV, and HR-HPV. In this analysis, any HPV refers to the presence of any of the 37 HPV genotypes. Infection with multiple types, defined as two or more HPV genotypes was also assessed compared to persons with one or no HPV infections. Demographic and sexual behavior characterstics by smoking status was compared using Pearson Chi-square test. In addition, difference in the prevalence of HPV genotypes by smoking status using Pearson Chi-square test was assessed. These analyses were stratified by MSM and MSW. To assess the association between HPV prevalence and smoking status, prevalence ratios (PR) and 95% confidence intervals (CI) using Poisson regression with robust variance estimation were calculated. Outcome variables for the models were: 1) Any HPV vs. No HPV, 2) HR-HPV vs. no HR-HPV, 3) LR-HPV vs. no LR-HPV, 4) multiple HPV infections vs no HPV/only one HPV infection. Known potential confounders were selected *a priori* to be used for adjustment in multivariale models. In the overall cohort, prevalence ratios were adjusted for age, country, sexual orientation (MSM or MSW) and lifetime female sexual partners. The MSW only model was adjusted for age, country, and lifetime female sexual partners. The MSM only model was adjusted for age, country, lifetime female and male sexual partners. All statistical tests were performed using STATA software, version 15.1 (StataCorp LLC, College Station, Texas).

## Results

A total of 1994 men were included in the analysis with 23% (n= 454) being current smokers, 19% (n=379) were former smokers and 58% (n=1161) having never smoked. The median pack-years smoked among current and former smokers was 3 years. **Table 1** shows the study population demographic characteristics stratified by smoking status. In this study population, 45% of participants were between 18-30 years old, 39% were between 31-44 years old and 15% were between 45-70 years old. Mexico had the highest proportion of participants that are current and former smokers (45% and 37% respectively). About 42% of MSW and 44% of MSM were ever smokers. Lifetime number of female sexual partners differed by smoking status (p<0.001), but was not significantly different for lifetime male sexual partners (p=0.64). Among current smokers, participants have been smoking for an average of 14 years with 9 cigarettes per day while former smokers had been smoking for an average of 11 years and 10 cigarettes per day.

**Table 1.** Demographic characteristic stratified by smoking status.

| **Table 1. Demographic characteristic stratified by smoking status** |  |  |  |  |
| --- | --- | --- | --- | --- |
| **Characteristic** | **Current**<br>**(n=454)** | **Former**<br>**(n=379)** | **Never**<br>**(n=1161)** | **P-value** |
| **Age categories** |  |  |  | <0.001 |
| 18-30 | 202 (44.5) | 105 (27.7) | 602 (51.8) |  |
| 31-44 | 193 (42.5) | 163 (43.0) | 431 (37.1) |  |
| 45-70 | 59 (13.0) | 111 (29.3) | 128 (11.0) |  |
| **Country of Birth** |  |  |  | <0.001 |
| USA | 110 (24.2) | 120 (31.7) | 396 (34.1) |  |
| Mexico | 205 (45.1) | 142 (37.5) | 304 (26.2) |  |
| Brazil | 139 (30.6) | 117 (30.9) | 461 (39.7) |  |
| **Race** |  |  |  | <0.001 |
| White | 170 (37.4) | 182 (48) | 562 (48.4) |  |
| Black | 63 (13.9) | 39 (10.3) | 204 (17.6) |  |
| Asian/PI | 6 (1.3) | 3 (0.8) | 32 (2.8) |  |
| Other | 205 (44.1) | 149 (39.3) | 328 (28.2) |  |
| Refused | 10 (2.2) | 6 (1.6) | 35 (3) |  |
| **Ethnicity** |  |  |  | <0.001 |
| Non Hispanic | 198 (43.6) | 186 (49.5) | 667 (58.4) |  |
| Hispanic | 256 (56.4) | 190 (50.5) | 476 (41.6) |  |
| **Marital status** |  |  |  | <0.001 |
| Divorced/separated | 52 (11.5) | 44 (11.6) | 88 (7.6) |  |
| Married/cohabitating | 205 (45.4) | 228 (60.2) | 497 (43.0) |  |
| Single | 195 (43.1) | 107 (28.2) | 572 (49.4) |  |
| **Sexual Orientation** |  |  |  | 0.05 |
| MSM | 97 (21.4) | 84 (22.2) | 224 (19.3) |  |
| MSW | 303 (66.7) | 267 (70.5) | 787 (67.8) |  |
| Unknown | 54 (11.9) | 28 (7.4) | 150 (12.9) |  |
| **Lifetime Male Sexual Partners** |  |  |  | 0.637 |
| 0-1 | 370(81.5) | 316(83.38) | 982(84.58) |  |
| 2-9 | 37(8.15) | 35(9.23) | 91(7.84) |  |
| 10 to 49 | 28(6.17) | 15(3.96) | 49(4.22) |  |
| 50+ | 11(2.42) | 9(2.37) | 28(2.41) |  |
| Missing | 8 (1.8) | 4 (1.1) | 11 (0.9) |  |
| **Lifetime Female Sexual Partners** |  |  |  | <0.001 |
| 0-1 | 62 (13.7) | 33 (8.7) | 292 (25.1) |  |
| 2-9 | 163 (36.0) | 141 (37.2) | 477 (41.1) |  |
| 10-49 | 159 (35.0) | 161 (42.5) | 297 (25.6) |  |
| 50+ | 37 (8.1) | 25 (6.6) | 50 (4.3) |  |
| Missing | 33 (7.3) | 19 (5.0) | 45 (3.9) |  |
| **Years smoked** |  |  |  |  |
| Mean (std) | 14.7 (11.3) | 10.8 (9.2) | n/a |  |
| Median (IQR) | 12 (7-20) | 9 (3-15) | n/a |  |
| **Average cigarettes per day** |  |  |  |  |
| Mean (std) | 8.5 (7.7) | 10.4 (11.7) | n/a |  |
| Median (IQR) | 6 (3-12) | 5 (2-15) | n/a |  |
| **Pack years** |  |  |  |  |
| Mean (std) | 7.5 (10.4) | 8.1 (15.7) | n/a |  |
| Median (IQR) | 3.6 (1.0-9.8) | 2.4 (0.5-10) |  |  |
| Abbreviations: std= standard deviation; IQR= interquartile range<br>P-values were calculated using chi square test |  |  |  |  |

Anal HPV genotype prevalence by smoking status is desribed in **Table 2**. Overall, prevalence of any anal HPV was 23.6% among current smoker, 15.8% among former smoker and 17.4% among never smoker (p-value =0.005). The prevalence of anal LR-HPV was highest in current smoker (17.1%) followed by former smoker (12.7%) and never smoker (11.5%) (p-value= 0.002). When stratified by sexual orientation, there were no significant differences in the prevalence of anal HPV groupings among MSM by smoking status. However, among MSW the prevalence of any anal HPV was 16.8% in current smoker, 9.7% among former smoker and 10.3% among never smoker (p-value=0.006). The prevalence of anal LR- and HR-HPV were also significantly higher among current MSW smokers compared to negative smokers. The prevalence of multiple infections was significantly higher among current smokers in both the MSM and overall groups. Prevalence of individual HPV genotypes by smoking status is shown in Supplemental Table 1. There was no significant differences in the prevalence of anal HPV by smoking intensity among current or former smokers (Supplemental Table 2).

**Table 2.**
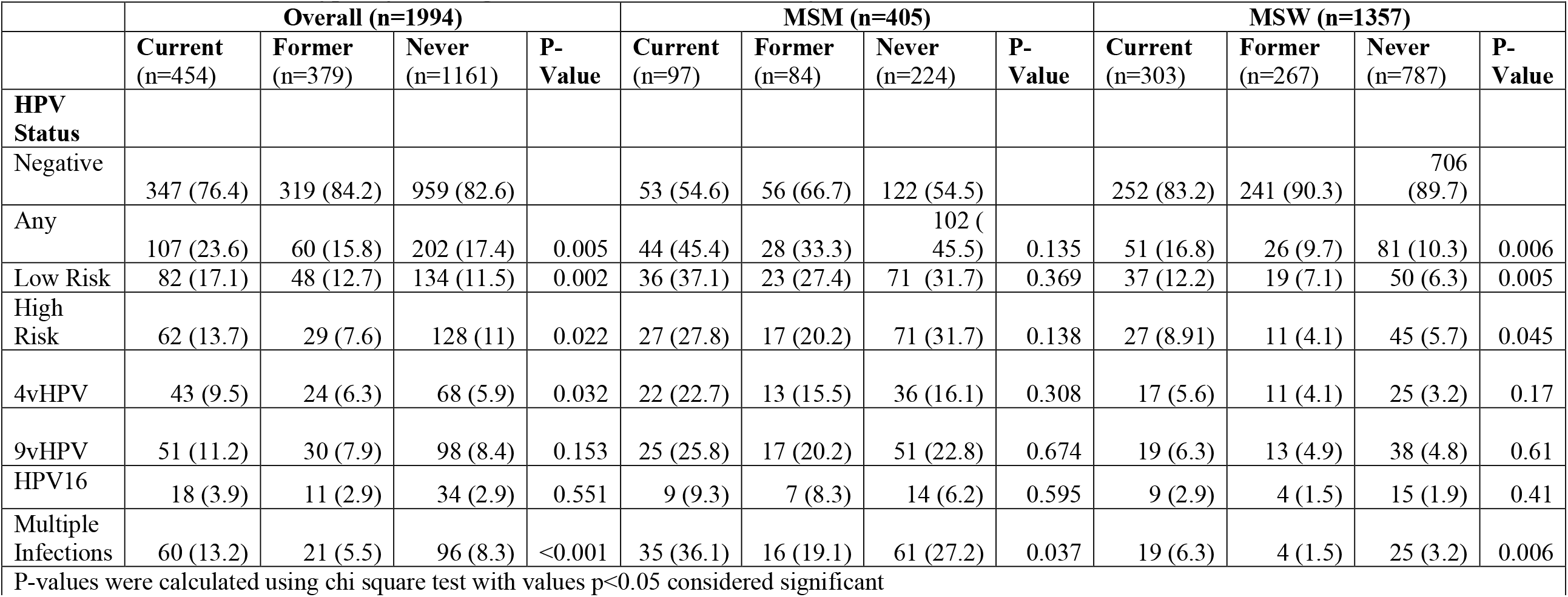
Prevalence of HPV types by smoking status overall and stratified by sexual orientation.

The association between smoking status and pack-years smoked and anal HPV infection overall is presented in **Table 3**. Current smokers had a higher prevalence of any anal HPV (adjusted PR (aPR)=1.36, 95%CI: 1.06-1.73) and LR-HPV (aPR=1.59, 95%CI: 1.20-2.12) compared to never smokers after adjusting for age, country, sexual orientation and lifetime female sexual partners. Current smokers had a significanly higher prevalence of multiple infections (aPR= 1.57, 95%CI: 1.12-2.18) compared to never smokers. There were no statisically significant difference in the prevalence of anal HPV between former smoker and never smokers. The association between smoking intensity, dichotomotized pack-years (median=3 pack-years) smoked among current and former smokers and less than 3 pack-years or greater than or equal to 3 pack-years, and the prevalence of anal HPV was assessed. There were no differences by HPV grouping, but we did find that current smokers with ≥3 pack-years had a significanlty higher prevalence of multiple infections (aPR= 1.63, 95%CI: 1.09-2.45) compared to never smokers. In a post hoc analysis, we stratified by country and found that current smokers in the US had a significanly higher prevalence of LR-HPV (aPR=3.25, 95% CI: 1.67-6.29) compared to never smokers (Supplemental Table 3). There were no significant differences in Brazil or Mexico comparing the prevalence of anal HPV by smoking status (Supplemental Table 4 and 5).

**Table 3:** Prevalence Ratios for all.

|  | Any HPV <sup>a</sup> |  | High Risk HPV <sup>a</sup> |  | Low Risk HPV <sup>a</sup> |  | Multiple Infections <sup>b</sup> |  |
| --- | --- | --- | --- | --- | --- | --- | --- | --- |
|  | uPR<br>(95%CI) <sup>c</sup> | aPR (95%<br>CI) <sup>d</sup> | uPR<br>(95%CI) <sup>c</sup> | aPR (95%<br>CI) <sup>d</sup> | uPR<br>(95%CI) <sup>c</sup> | aPR (95%<br>CI) <sup>d</sup> | uPR<br>(95%CI) <sup>c</sup> | aPR (95%<br>CI) <sup>d</sup> |
| **Smoking Status** |  |  |  |  |  |  |  |  |
| Never | 1.0(ref) | 1.0 (ref) | 1.0(ref) | 1.0 (ref) | 1.0(ref) | 1.0 (ref) | 1.0(ref) | 1.0(ref) |
| Former | 0.91 (0.68-1.21) | 0.97 (0.72-1.31) | 0.69 (0.46-1.04) | 0.77 (0.51-1.17) | 1.09 (0.79-1.52) | 1.21 (0.86-1.71) | 0.67(0.42-1.07) | 0.78(0.48-1.28) |
| Current | 1.35 (1.07-1.71) | 1.36 (1.06-1.73) | 1.23 (0.91-1.68) | 1.24 (0.91-1.70) | 1.56 (1.18-2.06) | 1.59 (1.20-2.12) | 1.59(1.15-2.20) | 1.57(1.12-2.18) |
| **Pack-years smoked** |  |  |  |  |  |  |  |  |
| Never | 1.0 (ref) | 1.0 (ref) | 1.0 (ref) | 1.0 (ref) | 1.0 (ref) | 1.0 (ref) | 1.0(ref) | 1.0(ref) |
| <3 pack-years (current<br>smoker) | 1.36 (0.99-1.86) | 1.41 (1.02-1.95) | 1.31 (0.88-1.96) | 1.32 (0.87-1.99) | 1.42 (0.97-2.07) | 1.51 (1.02-2.23) | 1.57(1.03-2.41) | 1.49(0.96-2.34) |
| ≥3 pack-years (current<br>smoker) | 1.35 (1.0-1.81) | 1.31 (0.97-1.78) | 1.17 (0.79-1.73) | 1.18 (0.79-1.77) | 1.68 (1.21-2.34) | 1.66 (1.18-2.34) | 1.61(1.08-2.39) | 1.63(1.09-2.45) |
| Never |  |  |  |  |  |  |  |  |
| <3 pack-years (former<br>smoker) | 1.02 (0.72-1.46) | 1.11 (0.77-1.59) | 0.89 (0.56-1.44) | 0.98 (0.61-1.59) | 1.20 (0.79-1.80) | 1.36 (0.89-2.07) | 0.89(0.52-1.55) | 1.01(0.58-1.76) |
| ≥3 pack-years (former<br>smoker) | 0.78 (0.51-1.19) | 0.80 (0.51-1.25) | 0.46 (0.23-0.91) | 0.51 (0.25-1.02) | 0.98 (0.61-1.57) | 1.04 (0.63-1.71) | 0.41(0.18-0.93) | 0.49(0.21-1.14) |

<sup>a</sup> Any, High Risk, Low Risk = models assessed the prevalence of one or more anal HPV infections for that grouping
<sup>b</sup> Multiple infections= models assessed having two or more prevalent any anal HPV infections
<sup>c</sup> aPR=unadjusted PR<sup>d</sup> aPR= multivariable PR adjusted for age, country, sexual orientation and lifetime female sexual partners

The association between smoking status and anal HPV infection stratified by sexual orientation is presented in **Tables 4 and 5**. There were no statistically significant differences in anal HPV prevalence by smoking status among MSM. Among MSW, current smokers had an increased prevalence of LR-HPV (aPR=1.60 95% CI: 1.02-2.50) compared to never smokers after adjusting for age, country, and lifetime female sexual partners. Current MSW smokers were more likely to have multiple HPV infections (aPR= 1.79, 95%CI: 1.35-2.39) compared to never smokers. Additonaly, when smoking intensity was dichotomized (<3 or ≥3 pack-years) among current smokers, both were associated with increased prevalence of mulitple infections. There were no differences between former and never smokers and anal HPV prevalence among MSW.

**Table 4.** HPV prevalence ratios by smoking status among MSM.

|  | Any HPV <sup>a</sup> |  | High Risk HPV <sup>a</sup> |  | Low Risk HPV <sup>a</sup> |  | Multiple Infections <sup>b</sup> |  |
| --- | --- | --- | --- | --- | --- | --- | --- | --- |
|  | uPR<br>(95%CI) <sup>c</sup> | aPR (95%<br>CI) <sup>d</sup> | uPR<br>(95%CI) <sup>c</sup> | aPR (95%<br>CI) <sup>d</sup> | uPR<br>(95%CI) <sup>c</sup> | aPR (95%<br>CI) <sup>d</sup> | uPR<br>(95%CI) <sup>c</sup> | aPR (95%<br>CI) <sup>d</sup> |
| **Smoking Status** |  |  |  |  |  |  |  |  |
| **Never** | 1.0 (ref) | 1.0 (ref) | 1.0 (ref) | 1.0 (ref) | 1.0 (ref) | 1.0 (ref) | 1.0(ref) | 1.0 (ref) |
| **Former** | 0.73 (0.48-1.11) | 0.93(0.6-1.45) | 0.64 (0.38-1.08) | 0.81(0.46-1.42) | 0.86 (0.54-1.38) | 1.08(0.66-1.77) | 0.71(0.54-0.93) | 0.92(0.69-1.24) |
| **Current** | 0.99 (0.69-1.42) | 0.93(0.65-1.35) | 0.88 (0.56-1.37) | 0.79(0.5-1.25) | 1.17 (0.78-1.75) | 1.1(0.73-1.66) | 1.04(0.83-1.31) | 0.94(0.75-1.19) |
| **Pack-years smoked** |  |  |  |  |  |  |  |  |
| **Never** | 1.0 (ref) | 1.0 (ref) | 1.0 (ref) | 1.0 (ref) | 1.0 (ref) | 1.0 (ref) | 1.0 (ref) | 1.0 (ref) |
| **<3 pack-years**<br>**(current smoker)** | 1.15 (0.71-1.87) | 0.87(0.53-1.44) | 0.99 (0.54-1.83) | 0.71(0.37-1.34) | 1.33 (0.77-2.28) | 0.99(0.56-1.75) | 1.29(0.96-1.75) | 0.91(0.66-1.24) |
| **≥3 pack-years**<br>**(current smoker)** | 0.89 (0.57-1.39) | 0.99(0.64-1.57) | 0.8 (0.46-1.39) | 0.88(0.5-1.55) | 1.07 (0.65-1.76) | 1.22(0.74-2.02) | 0.88(0.66-1.18) | 0.99(0.74-1.34) |
| **Never** | 1.0 (ref) | 1.0 (ref) | 1.0 (ref) | 1.0 (ref) | 1.0 (ref) | 1.0 (ref) | 1.0(ref) | 1.0(ref) |
| **<3 pack-years**<br>**(former smoker)** | 1.01 (0.61-1.67) | 1.14(0.68-1.90) | 0.89 (0.47-1.68) | 0.96(0.50-1.85) | 1.29 (0.75-2.23) | 1.41(0.81-2.46) | 1.11(0.81-1.53) | 1.23(0.89-1.70) |
| **≥3 pack-years**<br>**(former smoker)** | 0.49 (0.25-0.93) | 0.69(0.34-1.37) | 0.42 (0.18-0.97) | 0.62(0.25-1.51) | 0.49 (0.22-1.07) | 0.67(0.3-1.54) | 0.36(0.22-0.58) | 0.53(0.31-0.88) |

<sup>a</sup> Any, High Risk, Low Risk = models assessed the prevalence of one or more anal HPV infections for that grouping
<sup>b</sup> Multiple infections= models assessed having two or more prevalent any anal HPV infections
<sup>c</sup> aPR=unadjusted PR<sup>d</sup> aPR= multivariable PR adjusted for age, country, sexual orientation lifetime female sexual partners, and lifetime male sexual partners

**Table 5.** HPV prevalence ratios by smoking status among MSW.

|  | Any HPV <sup>a</sup> |  | High Risk HPV <sup>a</sup> |  | Low Risk HPV <sup>a</sup> |  | Multiple Infections <sup>b</sup> |  |
| --- | --- | --- | --- | --- | --- | --- | --- | --- |
|  | uPR<br>(95%CI) <sup>c</sup> | aPR (95%<br>CI) <sup>d</sup> | uPR<br>(95%CI) <sup>c</sup> | aPR (95%<br>CI) <sup>d</sup> | uPR<br>(95%CI) <sup>c</sup> | aPR (95%<br>CI) <sup>d</sup> | uPR<br>(95%CI) <sup>c</sup> | aPR (95%<br>CI) <sup>d</sup> |
| **Smoking Status** |  |  |  |  |  |  |  |  |
| **Never** | 1.0 (ref) | 1.0 (ref) | 1.0 (ref) | 1.0 (ref) | 1.0 (ref) | 1.0 (ref) | 1.0(ref) | 1.0 (ref) |
| **Former** | 0.95 (0.61-1.47) | 0.81 (0.51-1.28) | 0.72 (0.37-1.39) | 0.68 (0.35-1.35) | 1.12 (0.66-1.89) | 0.94 (0.54-1.63) | 0.83(0.57-1.22) | 0.77(0.52-1.14) |
| **Current** | 1.63 (1.15-2.32) | 1.39 (0.96-2.02) | 1.56 (0.97-2.51) | 1.41 (0.85-2.33) | 1.92 (1.25-2.94) | 1.60 (1.02-2.50) | 1.97(1.5-2.59) | 1.79(1.35-2.39) |
| **Pack-years smoked** |  |  |  |  |  |  |  |  |
| **Never** | 1.0 (ref) | 1.0 (ref) | 1.0 (ref) | 1.0 (ref) | 1.0 (ref) | 1.0 (ref) | 1.0 (ref) | 1.0 (ref) |
| **<3 pack-years (current smoker)** | 1.71 (1.09-2.65) | 1.55 (0.96-2.48) | 1.77 (0.99-3.18) | 1.64 (0.87-3.06) | 1.7 (0.97-2.99) | 1.48 (0.81-2.69) | 1.77(1.24-2.53) | 1.68(1.15-2.46) |
| **≥3 pack-years (current smoker)** | 1.56 (1-2.45) | 1.27 (0.79-2.03) | 1.35 (0.72-2.56) | 1.22 (0.63-2.38) | 2.13 (1.28-3.55) | 1.69 (0.99-2.89) | 2.16(1.55-2.99) | 1.90(1.34-2.69) |
| **Never** | 1.0 (ref) | 1.0 (ref) | 1.0 (ref) | 1.0 (ref) | 1.0 (ref) | 1.0 (ref) | 1.0(ref) | 1.0(ref) |
| **<3 pack-years (former smoker)** | 0.97 (0.56-1.68) | 0.86 (0.49-1.51) | 1.05 (0.51-2.15) | 0.99 (0.47-2.06) | 0.94 (0.46-1.92) | 0.82 (0.40-1.69) | 0.96(0.61-1.51) | 0.92(0.58-1.46) |
| **≥3 pack-years (former smoker)** | 0.91 (0.48-1.71) | 0.73 (0.38-1.42) | 0.29 (0.07-1.23) | 0.27 (0.06-1.15) | 1.34 (0.68-2.65) | 1.10 (0.54-2.27) | 0.67(0.37-1.22) | 0.59(0.32-1.1) |

<sup>a</sup> Any, High Risk, Low Risk = models assessed the prevalence of one or more anal HPV infections for that grouping
<sup>b</sup> Multiple infections= models assessed having two or more prevalent any anal HPV infections
<sup>c</sup> aPR=unadjusted PR
<sup>d</sup> aPR= mutivariable PR adjusted for age, country, sexual orientation lifetime female sexual partners

## Discussion

In this multinational cohort of 1994 HIV-negative men, the prevalence of anal HPV by smoking status and smoking intensity was assessed. Current smokers had a significantly higher prevalence of any anal HPV and LR-HPV compared to never smokers. Additionally, current smokers were more likely to have multiple HPV infections than never smokers. There were no differences in anal HPV prevalence between former and never smokers. When models were stratified by sexual orientation, MSW that currently smoke had a significantly higher prevalence of anal LR-HPV and multiple HPV infections compared to never smokers after adjusted for age, country, and female sexual partners. Smoking status was not associated with anal HPV prevalence among MSM.

The prevalence of anal HPV among MSM without HIV is common with 47% being infected with any HPV in a recent meta-analysis.^15^ In this study, 43% of MSM without HIV were infected with any HPV. Results from this study found that HPV prevalence was not associated with smoking status among HIV-negative MSM. Among MSM without HIV from Toronto, current smoking was not associated with the prevalence of any HPV or HR-HPV compared to those not currently smoking.^16^ Among MSM without HIV participating in the EXPLORE study, smoking was not associated with the prevalence of HPV.^17^ We previously reported risk factors associated with anal HPV prevalent among MSM in the *HIM Study*, and in adjusted models found no association between smoking and prevalent anal HPV infection.^18^ These results are consistent with other studies. While Goldstone, et al reported a significant increased prevalence of any HPV among former smokers compared to never smokers but no difference between current and never smokers.^19^ Smoking may not be associated with HPV infection; it could be associated with HPV persistence and disease development in MSM given that smoking is associated with anal cancer risk.

To the best of our knowledge, we are the first to report the role of smoking intensity and duration in anal HPV prevalence among MSW without HIV. We previously reported risk factors associated with anal HPV prevalent among MSW in the *HIM Study*, and in adjusted models found that formers smokers were significanly less likley to have a prevalent HPV infection than never smokers.^18^ Among MSW with HIV, current smokers were significanly more likely to have prevalent HPV infection compared to nonsmokers.^20^ In this study, the odds of any HPV were 1.70 times higher (95% CI: 1.17-2.52) in current smokers compared to never smokers. Current smokers also had increased prevalence of multiple infections compared to never smokers. Multiple anal HPV infections have been previously shown to be a risk factor anal pre-cancer. ^21, 22^ MSW are likley not engaging in receptive anal intercourse, yet we are detecting HPV within the anal canal. It is possible that these infections are occuring through autoinnoculation or partner innoculation. Within the *HIM Study*, we have reported that MSW with a prior genital HPV infection were at a higher risk for a subsequent type-specific anal infection compared to men without genital infections.^23^ Current smokers are also more likely to have prevalent genital HPV infection compared to never smokers.^10^ Among MSW, smoking may impact the prevalence of anal HPV and more research is needed to determine the role of smoking in anal HPV persistence among MSW.

Tobacco smoking has been shown to be associated with anal cancer development in several studies. ^24, 25,26,27^ Several case-controls studies in both men and women in the general population have reported a two-fold increased odds of anal cancer among smokers compared to non-smokers. Similarly, studies among people living with HIV also found an association between smoking and anal cancer.^28^ Smoking has also been shown to increase the risk for other HPV-associated cancers. The tobacco use could be affecting HPV persistence or progression of HPV-related lesions to cancer by reducing the immune response to HPV infections. While MSM are at an increased risk for anal cancer, we found that the prevalence of anal HPV did not differ by smoking status. Anal HPV is very common among MSM, factors associated with anal HPV persistence and lesion development may be modified by smoking.

This study has several strengths including the large sample size, data collection from three international clinical sites and ability to assess the association among both MSW and MSM. Very few studies have assessed anal HPV prevalence among MSW populations or MSM without HIV. Data collection and specimen processing was consistent across the three clinical sites. In order to interpret our results, it is important to keep in mind some of the limitations of this study. The HIM Study is not a populaiton-based study, but the demographics and age, ethnicity and education of the men included at each site are similar to the underlying population within their respective communities. However, our results may not be generalizable to the broader male population within these communities. For this analysis, we only assessed the association between smoking and baseline HPV infections. It would be interesting to assess the relationship between smoking and anal HPV incidence and clearance. Lifetime number of sexual partners were adjusted for the models, but this does not capture the different types of sexual intercourse. There are differences in anal HPV prevalence among MSM by type of anal intercourse, i.e., either receptive or insertive. Further research is needed to understand fully the association between smoking and anal HPV in men. It is particularly important to study this association in both MSM and MSW.

In conclusion, our study found currently smoking was associated with a higher prevalence of anal HPV. While we did not see a difference in anal HPV prevalence among MSM by smoking status, MSW that currently smoked had a higher prevalence of LR-HPV. Smoking may alter the immune response to HPV infection. Therefore, it is important to conduct more studies on the effects of smoking on anal HPV persistence and educate the public about smoking as a risk factor for anal HPV infection and anal cancer.

## Data Availability

All data produced in the present study are available upon reasonable request to the authors.

## Acknowledgements

This research was supported in part by research funding from Merck Sharp & Dohme Corp. The opinions expressed in this paper are those of the authors and do not necessarily represent those of Merck Sharp & Dohme Corp.

We thank the HIM Study teams and participants in the United States (Moffitt Cancer Center, Tampa), Brazil (Centro de Referência e Treinamento em DST/AIDS, Fundação Faculdade de Medicina Instituto do Câncer do Estado de São Paulo, Ludwig Institute for Cancer Research, São Paulo), and Mexico (Instituto Mexicano del Seguro Social, Instituto Nacional de Salud Pública, Cuernavaca).

