## Supplemental tables for "The association between smoking and anal human papillomavirus in the HPV infection in Men Study"

| **Supplemental Table 1. Prevalence of HPV types by smoking status** | | | | |
| --- | --- | --- | --- | --- |
| **HPV** | **Current (n=454)** | **Former (n=379)** | **Never (n=1161)** | **P-value** |
| HR |  |  |  |  |
| 16 | 18 (4) | 11 (2.9) | 34 (2.9) | 0.54 |
| 18 | 8 (1.8) | 3 (0.8) | 11 (0.9) | 0.30 |
| 31 | 2 (0.4) | 2 (0.5) | 5 (0.4) | 0.97 |
| 33 | 1 (0.2) | 1 (0.3) | 7 (0.6) | 0.49 |
| 35 | 4 (0.9) | 3 (0.8) | 6 (0.5) | 0.67 |
| 39 | 2 (0. 4) | 2 (0.5) | 14 (1.2) | 0.24 |
| 45 | 7 (1.5) | 4 (1.1) | 12 (1) | 0.68 |
| 51 | 12 (2.6) | 7 (1.9) | 23 (2) | 0.66 |
| 52 | 4 (0.9) | 3 (0.8) | 8 (0.7) | 0.92 |
| 56 | 5 (1.1) | 2 (0.5) | 12 (1) | 0.63 |
| 58 | 4 (0.9) | 3 (0.8) | 9 (0.9) | 0.98 |
| 59 | 6 (1.3) | 3 (0.8) | 20 (1.7) | 0.41 |
| 68 | 7 (1.5) | 2 (0.5) | 9 (0.8) | 0.24 |
| LR |  |  |  |  |
| 6 | 16 (3.5) | 9 (2.4) | 29 (2.5) | 0.47 |
| 11 | 5 (1.1) | 1 (0.3) | 4 (0.3) | 0.12 |
| 26 | 0 | 0 | 1(0.1) | 0.7 |
| 40 | 5 (1.1) | 1 (0.3) | 2 (0.2) | 0.03 |
| 42 | 1 (0.2) | 0 | 5 (0.4) | 0.39 |
| 53 | 12 (2.6) | 6 (1.6) | 22 (1.9) | 0.51 |
| 54 | 7 (1.5) | 1 (0.3) | 9 (0.8) | 0.12 |
| 55 | 3 (0.7) | 2 (0.5) | 6 (0.5) | 0.94 |
| 61 | 10 (2.2) | 5 (1.3) | 16 (1.4) | 0.44 |
| 62 | 5 (1.1) | 9 (2.4) | 20 (1.7) | 0.37 |
| 64 | 0 | 0 | 0 |  |
| 66 | 7 (1.5) | 2 (0.5) | 9 (0.8) | 0.24 |
| 67 | 2 (0.4) | 0 | 1 (0.1) | 0.18 |
| 69 | 0 | 1 (0.3) | 1 (0.1) | 0.48 |
| 70 | 8 (1.8) | 2 (0.5) | 6 (0.5) | 0.03 |
| 71 | 5 (1.1) | 0 | 9 (0.8) | 0.15 |
| 72 | 3 (0.7) | 3 (0.8) | 5 (0.4) | 0.67 |
| 73 | 6 (1.3) | 3 (0.8) | 9 (0.8) | 0.56 |
| 81 | 10 (2.2) | 4 (1.1) | 11 (0.9) | 0.12 |
| 82 | 2 (0.4) | 1 (0.3) | 3 (0.3) | 0.83 |
| (139) 82s^b^ | 3 (0.7) | 0 | 1 (0.1) | 0.04 |
| 83 | 8 (1.8) | 1 (0.3) | 8 (0.7) | 0.04 |

| **Supplemental Table 2. Prevalence of HPV infection by smoking status and tertiles of smoking characteristics** | | | | | | | | |
| --- | --- | --- | --- | --- | --- | --- | --- | --- |
|  | Former | | | | Current | | | |
|  | T1 | T2 | T3 | P-value | T1 | T2 | T3 | P-value |
| **Tertiles smoked/day** | (n=131) | (n=121) | (n=127) |  | (n=155) | (n=180) | (n=119) |  |
| **HPV status** |  |  |  |  |  |  |  |  |
| Negative | 109 (83.2) | 102 (84.3) | 108 (85) |  | 121 (78.1) | 141 (78.3) | 85 (71.4) |  |
| Any HPV | 22 (16.8) | 19 (15.7) | 19 (15) | 0.92 | 34 (21.9) | 39 (21.7) | 34 (28.6) | 0.33 |
| High Risk | 13 (9.9) | 9 (7.4) | 7 (5.5) | 0.41 | 17 (11) | 25 (13.9) | 20 (16.8) | 0.37 |
| Low Risk | 16 (12.2) | 15 (12.4) | 17 (13.4) | 0.95 | 25 (16.1) | 29 (16.1) | 28 (23.5) | 0.20 |
| **Tertiles of years smoked** | n=171 | n=122 | n=86 |  | n=134 | n=146 | n=174 |  |
| Negative | 145 (84.8) | 100 (82) | 74 (86) |  | 100 (74.6) | 115 (78.8) | 132 (75.9) |  |
| Any HPV | 26 (15.2) | 22 (18) | 12 (13.9) | 0.70 | 34 (25.4) | 31 (21.2) | 42 (24.1) | 0.70 |
| High Risk | 12 (7) | 11 (9) | 6(7) | 0.79 | 23 (17.2) | 17 (11.6) | 22 (12.6) | 0.36 |
| Low Risk | 21 (12.3) | 18 (14.7) | 9 (10.5) | 0.64 | 23 (17.2) | 23 (15.7) | 36 (20.7) | 0.49 |
| **Tertiles of pack years** | n=154 | n=107 | n=118 |  | n=136 | n=164 | n=154 |  |
| Negative | 125 (81.2) | 94 (87.8) | 100 (84.7) |  | 106 (77.9) | 126 (76.8) | 115 (74.7) |  |
| Any HPV | 29 (18.8) | 13 (12.1) | 18 (15.2) | 0.34 | 30 (22.1) | 38 (23.2) | 39 (25.3) | 0.80 |
| High Risk | 16 (10.4) | 6 (5.6) | 7 (5.9) | 0.25 | 21 (15.4) | 18 (11) | 23 (14.9) | 0.45 |
| Low Risk | 22 (14.3) | 10 (9.3) | 16 (13.6) | 0.47 | 22 (16.2) | 27 (16.5) | 33 (21.4) | 0.41 |
| P-values were calculated using chi square test comparing any HPV with no HPV, low-risk HPV with no low-risk HPV, High-risk HPV with no high-risk HPV group  Tertiles were defined by overall smoking characteristic distributions by smoking status | | | | | | | | |

| **Supplemental Table 3: Prevalence Ratios for the US** | | | | | | | | |
| --- | --- | --- | --- | --- | --- | --- | --- | --- |
|  | **Any HPV ^a^** | | **High Risk HPV ^a^** | | **Low Risk HPV ^a^** | | **Multiple Infections ^b^** | |
|  | **uPR (95%CI) ^c^** | **aPR (95% CI) ^d^** | **uPR (95%CI) ^c^** | **aPR (95% CI) ^d^** | **uPR (95%CI) ^c^** | **aPR (95% CI) ^d^** | **uPR (95%CI) ^c^** | **aPR (95% CI) ^d^** |
| **Smoking Status** |  |  |  |  |  |  |  |  |
| **Never** | 1.0(ref) | 1.0 (ref) | 1.0(ref) | 1.0 (ref) | 1.0(ref) | 1.0 (ref) | 1.0 (ref) | 1.0 (ref) |
| **Former** | 1.5 (0.89-2.52) | 1.27(0.72-2.26) | 1.1(0.88-3.01) | 1(0.47-2.13) | 2.2(1.11-4.32) | 1.75(0.82-3.73) | 1.17(0.43-3.27) | 0.99(0.31-3.15) |
| **Current** | 2.19(1.37-3.5) | 2.04(1.24-3.36) | 1.64(0.55-2.17) | 1.51(0.47-2.13) | 3.43(1.86-6.32) | 3.25(1.67-6.29) | 2.83(1.28-6.23) | 2.63(1.11-6.27) |
| **Pack-years smoked** |  |  |  |  |  |  |  |  |
| **Never** | 1.0 (ref) | 1.0 (ref) | 1.0 (ref) | 1.0 (ref) | 1.0 (ref) | 1.0 (ref) | 1.0 (ref) | 1.0 (ref) |
| **<3 pack-years (current smoker)** | 2.33(1.17-4.61) | 2.42(1.21-4.83) | 2.27(1.0-5.13) | 2.21(0.96-5.06) | 3.06(1.23-7.57) | 3.2(1.28-7.99) | 3.06(1.01-9.29) | 2.76(0.89-8.54) |
| **≥3 pack-years (current smoker)** | 2.12(1.23-3.66) | 1.78(0.97-3.28) | 1.31(0.61-2.85) | 1.06(0.45-2.49) | 3.62(1.84-7.11) | 3.22(1.46-7.07) | 2.71(1.09-6.72) | 2.43(0.80-7.35) |
| **Never** | 1.0(ref) | 1.0(ref) | 1.0(ref) | 1.0(ref) | 1.0(ref) | 1.0(ref) | 1.0 (ref) | 1.0 (ref) |
| **<3 pack-years (former smoker)** | 1.84(0.97-3.48) | 1.77(0.91-3.45) | 1.71(0.79-3.71) | 1.77(0.78-4) | 2.36(1.0-5.54) | 2.25(0.92-5.51) | 2.02(0.66-6.14) | 2.36(0.69-8.01) |
| **≥3 pack-years (former smoker)** | 1.21(0.59-2.47) | 0.84(0.38-1.84) | 0.56(0.17-1.83) | 0.39(0.11-1.36) | 2.06(0.88-4.85) | 1.37(0.52-3.61) | 0.44(0.06-3.36) | 0.25(0.03-2.15) |
| ^a^ Any, High Risk, Low Risk = models assessed the prevalence of one or more anal HPV infections for that grouping  ^b^ Multiple infections= models assessed having two or more prevalent any anal HPV infections  ^c^ aPR=unadjusted PR  ^d^ aPR= mutivariable PR adjusted for age, sexual orientation and lifetime female sexual partners | | | | | | | | |

| **Supplemental Table 4: Prevalence Ratios for Brazil** | | | | | | | | |
| --- | --- | --- | --- | --- | --- | --- | --- | --- |
|  | **Any HPV ^a^** | | **High Risk HPV ^a^** | | **Low Risk HPV ^a^** | | **Multiple Infections ^b^** | |
|  | **uPR (95%CI) ^c^** | **aPR (95% CI) ^d^** | **uPR (95%CI) ^c^** | **aPR (95% CI) ^d^** | **uPR (95%CI) ^c^** | **aPR (95% CI) ^d^** | **uPR (95%CI) ^c^** | **aPR (95% CI) ^d^** |
| **Smoking Status** |  |  |  |  |  |  |  |  |
| **Never** | 1.0(ref) | 1.0 (ref) | 1.0(ref) | 1.0 (ref) | 1.0(ref) | 1.0 (ref) | 1.0 (ref) | 1.0 (ref) |
| **Former** | 0.91(0.59-1.41) | 0.99(0.62-1.55) | 0.72(0.38-1.32) | 0.77(0.40-1.47) | 1.03(0.64-1.65) | 1.09(0.67-1.80) | 0.75(0.40-1.39) | 0.77(0.41-1.48) |
| **Current** | 1.29(0.9-1.84) | 1.19(0.83-1.72) | 1.31(0.83-2.06) | 1.19(0.75-1.88) | 1.3(0.87-1.95) | 1.19(0.79-1.80) | 1.42(0.90-2.23) | 1.27(0.8-2.0) |
| **Pack-years smoked** |  |  |  |  |  |  |  |  |
| **Never** | 1.0 (ref) | 1.0 (ref) | 1.0 (ref) | 1.0 (ref) | 1.0(ref) | 1.0 (ref) | 1.0 (ref) | 1.0 (ref) |
| **<3 pack-years (current smoker)** | 1.42(0.83-2.44) | 1.09(0.63-1.91) | 1.24(0.59-2.59) | 0.87(0.41-1.84) | 1.34(0.71-2.51) | 1.04(0.55-1.98) | 1.63(0.83-3.17) | 1.18(0.59-2.34) |
| **≥3 pack-years (current smoker)** | 1.23(0.8-1.87) | 1.26(0.82-1.94) | 1.33(0.79-2.52) | 1.41(0.83-2.41) | 1.28(0.8-2.05) | 1.29(0.79-2.09) | 1.32(0.77-2.26) | 1.33(0.76-2.29) |
| **Never** | 1.0(ref) | 1.0(ref) | 1.0(ref) | 1.0(ref) | 1.0(ref) | 1.0(ref) | 1.0 (ref) | 1.0 (ref) |
| **<3 pack-years (former smoker)** | 1.22(0.67-2.21) | 1.06(0.58-1.96) | 0.99(0.43-2.30) | 0.82(0.35-1.93) | 1.43(0.76-2.69) | 1.25(0.66-2.38) | 1.22(0.56-2.66) | 0.97(0.44-2.16) |
| **≥3 pack-years (former smoker)** | 0.74(0.42-1.31) | 0.91(0.49-1.67) | 0.55(0.24-1.28) | 0.74(0.31-1.77) | 0.8(0.43-1.51) | 0.98(0.50-1.89) | 0.49(0.19-1.21) | 0.59(0.23-1.54) |
| ^a^ Any, High Risk, Low Risk = models assessed the prevalence of one or more anal HPV infections for that grouping  ^b^ Multiple infections= models assessed having two or more prevalent any anal HPV infections  ^c^ aPR=unadjusted PR  ^d^ aPR= mutivariable PR adjusted for age, sexual orientation and lifetime female sexual partners | | | | | | | | |

| **Supplemental Table 5: Prevalence Ratios for Mexico** | | | | | | | | |
| --- | --- | --- | --- | --- | --- | --- | --- | --- |
|  | **Any HPV ^a^** | | **High Risk HPV ^a^** | | **Low Risk HPV ^a^** | | **Multiple Infections ^b^** | |
|  | **uPR (95%CI) ^c^** | **aPR (95% CI) ^d^** | **uPR (95%CI) ^c^** | **aPR (95% CI) ^d^** | **uPR (95%CI) ^c^** | **aPR (95% CI) ^d^** | **uPR (95%CI) ^c^** | **aPR (95% CI) ^d^** |
| **Smoking Status** |  |  |  |  |  |  |  |  |
| **Never** | 1.0(ref) | 1.0 (ref) | 1.0(ref) | 1.0 (ref) | 1.0(ref) | 1.0 (ref) | 1.0 (ref) | 1.0 (ref) |
| **Former** | 0.62(0.34-1.13) | 0.63(0.34-1.15) | 0.44(0.18-1.06) | 0.48(0.19-1.17) | 0.88(0.45-1.73) | 1.44(0.85-2.45) | 0.45(0.15-1.32) | 0.56(0.18-1.66) |
| **Current** | 1.14(0.74-1.75) | 1.05(0.67-1.62) | 1.07(0.61-1.88) | 0.93(0.52-1.66) | 1.48(0.88-2.48) | 0.93(0.47-1.85) | 1.72(0.93-3.17) | 1.6(0.85-3.04) |
| **Pack-years smoked** |  |  |  |  |  |  |  |  |
| **Never** | 1.0 (ref) | 1.0 (ref) | 1.0 (ref) | 1.0 (ref) | 1.0(ref) | 1.0 (ref) | 1.0 (ref) | 1.0 (ref) |
| **<3 pack-years (current smoker)** | 1.22(0.744-1.98) | 1.09(0.65-1.81) | 1.26(0.67-2.34) | 0.99(0.52-1.91) | 1.42(0.78-2.59) | 1.35(0.72-2.51) | 1.66(0.82-3.37) | 1.39(0.67-2.92) |
| **≥3 pack-years (current smoker)** | 1.03(0.56-1.89) | 0.98(0.52-1.83) | 0.78(0.33-1.89) | 0.81(0.33-1.98) | 1.57(0.8-3.08) | 1.59(0.79-3.18) | 1.79(0.81-3.98) | 2.02(0.88-4.61) |
| **Never** | 1.0(ref) | 1.0(ref) | 1.0(ref) | 1.0(ref) | 1.0(ref) | 1.0(ref) | 1.0 (ref) | 1.0 (ref) |
| **<3 pack-years (former smoker)** | 0.73(0.38-1.37) | 0.71(0.37-1.34) | 0.6(0.25-1.45) | 0.62(0.25-1.5) | 1.0(0.49-2.07) | 1.00(0.48-2.09) | 0.61(0.21-1.81) | 0.68(0.23-2.03) |
| **≥3 pack-years (former smoker)** | 0.33(0.81-1.37) | 0.36(0.08-1.55) | n/a | n/a | 0.55(0.13-2.31) | 0.67(0.15-2.96) | n/a | n/a |
| ^a^ Any, High Risk, Low Risk = models assessed the prevalence of one or more anal HPV infections for that grouping  ^b^ Multiple infections= models assessed having two or more prevalent any anal HPV infections  ^c^ aPR=unadjusted PR  ^d^ aPR= mutivariable PR adjusted for age, sexual orientation and lifetime female sexual partners | | | | | | | | |
